# Association of ICH Score with Withdrawal of Life-Sustaining Treatment: A Decade from the Florida Stroke Registry

**DOI:** 10.1101/2025.01.30.25321442

**Authors:** Nina Massad, Lili Zhou, Brian Manolovitz, Negar Asdaghi, Hannah Gardener, Hao Ying, Carolina M Gutierrez, Angus Jameson, David Rose, Mohan Kottapally, Amedeo Merenda, Kristine O’Phelan, Sebastian Koch, Jose G. Romano, Tatjana Rundek, Ayham Alkhachroum

## Abstract

**Background and Purpose:** The intracerebral hemorrhage (ICH) score was created as a tool improve communication and consistency among providers, and authors initially cautioned against its use as a predictor of outcomes. We aimed to investigate the association of ICH score with mortality and withdrawal of life-sustaining treatment (WLST).

**Methods:** Patients with a diagnosis of ICH were identified using data from Florida Stroke Registry (FSR) hospitals participating in the American Heart Association (AHA) Get with the Guidelines-Stroke (GWTG-S) from 2013-2022. Outcomes of WLST and in-hospital mortality were collected. ICH score was stratified into three groups: ICH score 0-2; 3-4; 5-6. Importance plots were generated to identify the most predictive factors associated with WLST. AUC-ROC curves were generated for logistic regression (LR) and random forest (RF) models, adjusted for relevant confounders. Secondary outcome analyses were performed using stratified univariate logistic regression to assess changes between 2015-2018 and 2019-2022.

**Results:** A total of 12,426 (26%) patients had documented ICH scores (mean age 69, 55% male, 56% white). The most predictive factors associated with WLST were ICH score, age, state region, presenting level of consciousness, insurance status and race (RF AUC=.94, LR AUC=.82). Mortality was 6.6%, 41.5% and 66% for ICH score 0-2, 3-4 and 5-6. Decision to WLST occurred more for ICH scores 3-4 (OR 9.35, 95% CI: 8.5-10.3) and ICH scores 5-6 (15.43, 95% CI: 15.28-22.74). Early WLST (< 2 days) was more likely for ICH score 3-4 (OR 2.97, 95% CI: 2.48-3.55) and score 5-6 groups (OR 9.51, 95% CI: 7.33-12.35).

**Conclusion:** Among ICH patients admitted across Florida, we noted a significant association between ICH score and likelihood of mortality, decision to WLST, and specifically WLST within two days of presentation. We identified the most predictive variable associated with WLST to be the ICH score. These findings suggest a continued influence of the self-fulfilling prophecy in ICH.

## INTRODUCTION

Intracerebral hemorrhage (ICH) has been consistently demonstrated to have higher morbidity and mortality risk than subarachnoid hemorrhage or ischemic stroke. ^1–3^ Decision to withdraw life-sustaining treatment (WLST) after ICH is common.^2,4–6^ Furthermore, we have previously demonstrated that mortality in ICH and other acute brain injury is largely mediated by WLST decision.^4,5^

Like other conditions within neurocritical care, ICH has faced an overemphasis and heavy reliance on prognostication scores.^7–9^ One such tool is the ICH score, a clinical grading scale developed both as an outcome risk stratification scale and as a tool for providers to standardize ICH severity and aid inter-provider communication.^10^ The score incorporates factors including age, hematoma volume, location, presence of intraventricular hemorrhage (IVH) and Glasgow coma scale (GCS), totaling a score from zero to six. When initially described, an association was demonstrated between higher ICH score and outcome, defined as 30-day mortality, however its use as a tool predictive of outcome was cautioned.

Our study aimed to investigate how initial ICH score value has impacted overall mortality, WLST and timing of WLST decision-making. We hypothesized that decision to WLST was highly associated with ICH score value. Additionally, we hypothesized an overall higher rate of early WLST with higher ICH score, an overall decrease in WLST independent of ICH score value, and a decrease in odds of mortality independent of ICH score value over the study period.

## METHODS

### Data Availability Statement

The Florida Stroke Registry (FSR) uses data from Get With the Guideline-Stroke (GWTG-S). As GWTG-S is collected primarily for quality improvement, data-sharing agreements require an application process for other researchers to access data. Research proposal can be submitted at www.heart.org/qualityresearch and will be considered by the GWTG-S and the FSR advisory and publication committees upon reasonable request.

### Study Population

The FSR is a comprehensive repository of data collected from 181 participating hospitals statewide; its aim is to identify disparities in stroke care and develop interventions to improve the quality of stroke care in Florida. The registry was initially funded by the National Institute of Health/National Institute of Neurological Disorders, then known as the Florida-Puerto Rico Collaboration to Reduce Stroke Disparities, and since 2017 has continued through funding support from the state of Florida (COHAN-A1).

This study was approved by the University of Miami’s institutional review board. Participating centers were given institutional ethics approval to enroll patients in the registry; centers were not required to obtain individual patient consent under the common rule or waiver of authorization and exception from subsequent review by their institutional review board. The study was performed in accordance with the ethical standards on human experimentation as established in the Helsinki Declaration of 1975.

Using the FSR, we identified eligible patients with a primary diagnosis of ICH from 2013 to 2022. Patients were identified based on recorded final clinical diagnosis of “Intracerebral hemorrhage” from Get with the Guidelines-Stroke (GWTG-S) questionnaire. From this pool, patients with documented ICH scores were further identified, and patients without listed ICH scores were excluded. Baseline characteristics were compared between patients with and without documented scores to assess for selection bias and ensure comparability between groups.

ICH scores were determined at initial presentation as a baseline severity score in initial evaluation of patients with ICH, the clinical grading scale as described in the initial literature describing the score (Hemphill Stroke). Elements of the score listed as follows: 1) Glasgow Coma Score or GCS (two points given to GCS 3-4, one point for GCS 5-12, and 0 points for GCS 13-15); 2) one point for Age ≥ 80; 3) one point for ICH volume ≥ 30mL; 4) one point for presence of intraventricular hemorrhage (IVH); 5) one point for infratentorial original of hemorrhage. As ICH score ranges from 0-6, scores were stratified into three groups: ICH score of 0 to 2; ICH score of 3 to 4 and ICH score of 5 to 6.

For patients identified to have a diagnosis of ICH, WLST decision during hospitalization was determined as a discrete data element based on having GWTG-S documented responses of WLST or transition to comfort measures only. Furthermore, early WLST was defined as occurring within the first 0 to 1 day after presentation, whereas late WLST occurred day 2 or more after presentation as per (GWTG-S) questionnaire.^2,4^

Data collected included demographic information (age, sex, race/ethnicity), insurance status (private, Medicare, Medicaid, self/no-insurance, and unknown), information regarding stroke center type (comprehensive stroke center, thrombectomy capable stroke center, or primary stroke center), comorbidities (hypertension, diabetes, obesity, prior history of smoking, alcohol/drug use, atrial fibrillation or flutter, coronary artery disease, peripheral vascular disease, prior stroke, heart failure), prior ambulation status, intravenous thrombolysis or endovascular treatment, and surgical treatment (craniotomy or evacuation, external ventricular drain, multiple procedures or other). Information regarding disease severity was also collected, specifically ICH and GCS scores. Information regarding the state of Florida region was divided into east center, west central, north, panhandle and south Florida. Information regarding in-hospital mortality, ambulation status on discharge (independent or needs assistance), and discharge disposition (home/rehab or other).

### Statistical Analysis

The primary goal of this study was to identify the extent to which ICH score among other factors was associated with the decision to WLST after ICH. The secondary goal was to identify potential temporal differences in associations between ICH score value and mortality, rates of decision to WLST, and rates of early WLST.

For patient characteristics, continuous variables were summarized as median with first and third quartiles (Q1 and Q3). Descriptive statistics were performed using Pearson chi-squared and Kruskall-Wallis tests.

Importance plots were generating using random forest (RF) for factors associated with WLST, and thereby providing a list of most significant variables by a mean decrease in Gini. The following factors were included in the importance plot: ICH score value, age, sex, race/ethnicity, insurance status, smoking, drug/alcohol use, hypertension, diabetes, obesity, atrial fibrillation/atrial flutter, coronary artery disease, peripheral vascular disease, prior stroke or TIA, prior ambulation, stroke center type, surgical treatment, state region, and impaired level of consciousness on presentation. The variables included were those with clinical relevance and low missingness of less than 5%.

After reviewing the importance plot, including the most predictive features of decision to WLST, logistic regression (LR) and RF cross validation was performed. We generated area under the curve (AUC) of the receiver operating curve (ROC) to evaluate performance of the LR and RF models, using 70/15/15 for training/testing/validation. Both models incorporated the top 10 variables identified as predictors of WLST decision, including ICH score, age, region, impaired level of consciousness on presentation, insurance status, race, surgical treatment type, sex, stroke center type and diabetes.

Analysis of secondary outcomes was performed using univariate logistic regression to evaluate the association between: ICH score and mortality; ICH score and overall WLST; ICH score and early WLST. Data from 2015 to 2022 were utilized for secondary outcome analysis, and ICH group 0-2 was used as the reference group for the analyses. Data from the 2015-2022 was utilized, estimated at 95% confidence intervals (CI). We then performed a stratified univariate logistic regression for two time periods: 2015-2018 and 2019-2022, assessing for any temporal differences between ICH score and secondary outcomes. Models were adjusted for age, sex, race/ethnicity, comorbidities (hypertension, diabetes, obesity, prior stroke or TIA), region, insurance status and hospital type.

Results were reported as odds ratios (ORs) with 95% CIs and corresponding p-values, with statistical significance set at p<0.05; analyses were performed using SAS Version 9.4 software (SAS Institute), MATLAB 2021b, and R (3.6.0).

## RESULTS

### Study population

A total of 60,729 patients with a diagnosis of ICH were included in the FSR between 2013 and 2022. Of these patients, we studied a total of 12,426 (20%) who had documented ICH scores. Intergroup differences between ICH patients with and without documented ICH scores are shown in the supplementary table. Patients with documented ICH scores had similar rates of in-hospital mortality overall compared to patients with ICH score data missing (17% with scores versus 18% without ICH scores).

Of the 12,426 patients with documented ICH scores included, median age was 69 (SD = 14.96); 45% were women, 56% were White, 24% were Black, 20% Hispanic. Of these patients, 76% had hypertension, 25% had diabetes, 24% had obesity, 23% had prior history of stroke or transient ischemic attack (TIA). Patients were seen primarily in comprehensive stroke centers (84%) and thrombectomy-capable stroke centers (5%); median time from onset of symptoms to arrival time was 177 minutes (61, 515). More detailed characteristic data stratified by ICH score group are shown in table 1.

**Table 1.**
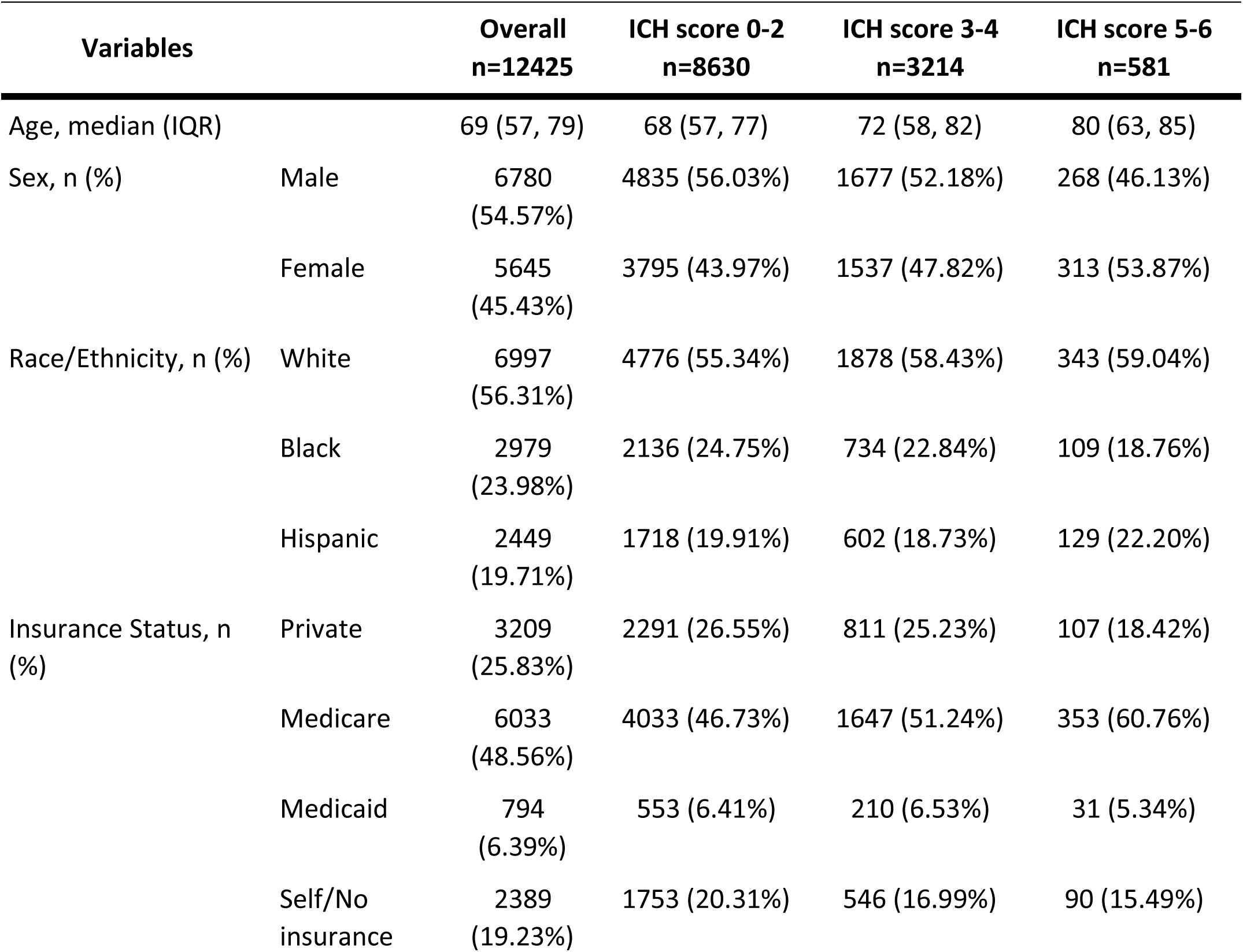
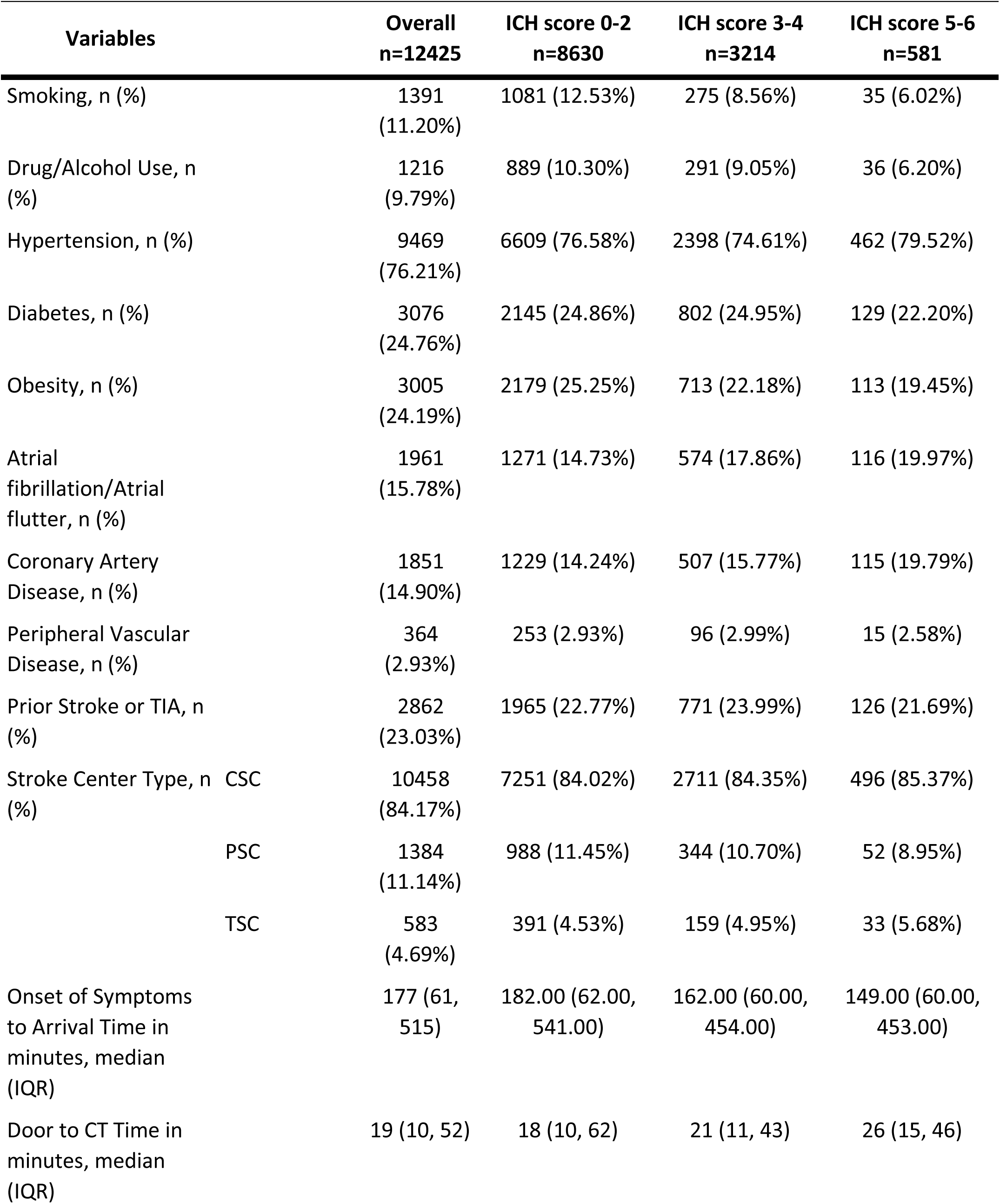
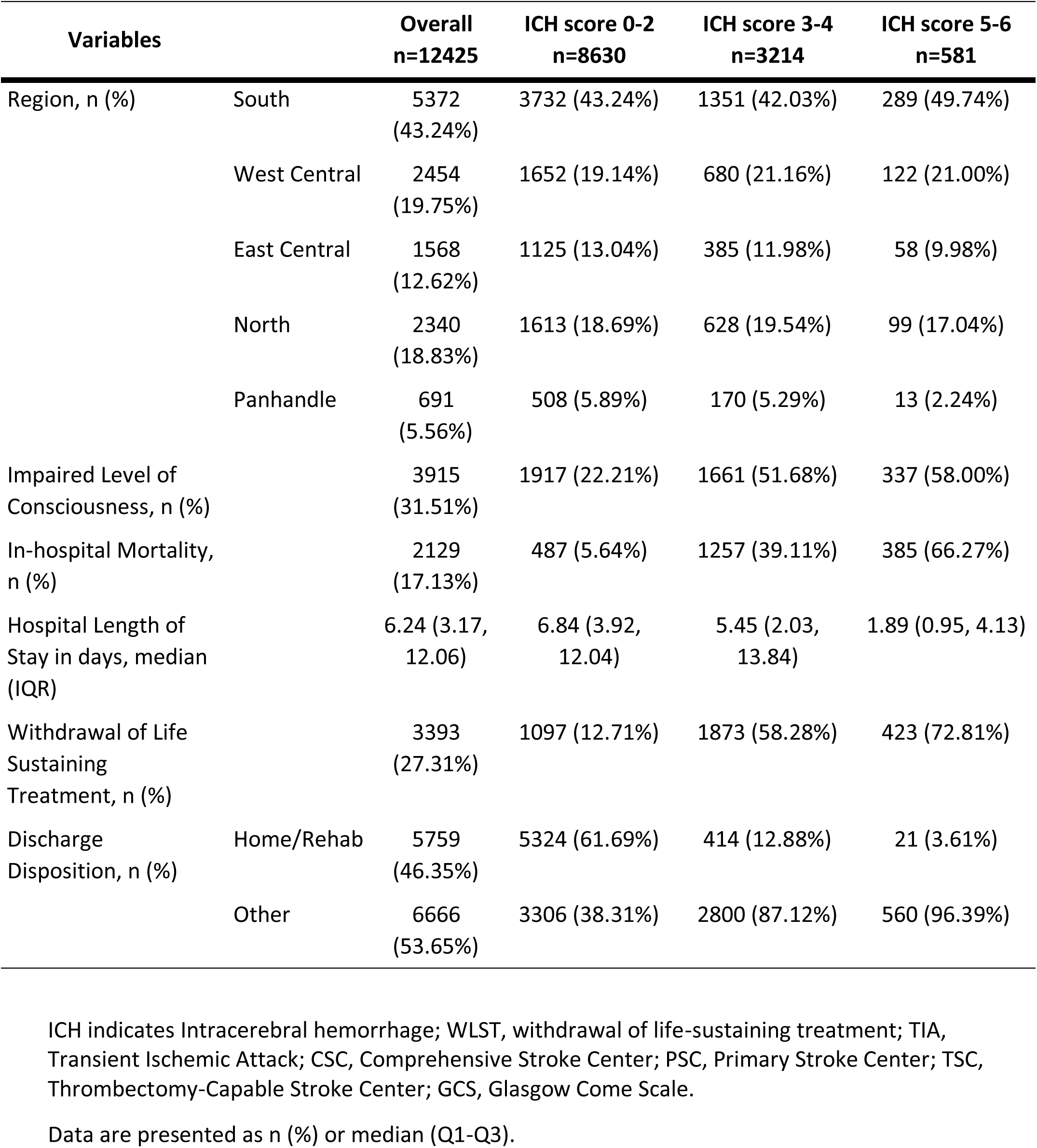
Baseline characteristics of ICH patients in FSR from 2013 to 2022 stratified by ICH score groups.

| Variables |  | Overall<br>n=12425 | ICH score 0-2<br>n=8630 | ICH score 3-4<br>n=3214 | ICH score 5-6<br>n=581 |
| --- | --- | --- | --- | --- | --- |
| Age, median (IQR) |  | 69 (57, 79) | 68 (57, 77) | 72 (58, 82) | 80 (63, 85) |
| Sex, n (%) | Male | 6780<br>(54.57%) | 4835 (56.03%) | 1677 (52.18%) | 268 (46.13%) |
|  | Female | 5645<br>(45.43%) | 3795 (43.97%) | 1537 (47.82%) | 313 (53.87%) |
| Race/Ethnicity, n (%) | White | 6997<br>(56.31%) | 4776 (55.34%) | 1878 (58.43%) | 343 (59.04%) |
|  | Black | 2979<br>(23.98%) | 2136 (24.75%) | 734 (22.84%) | 109 (18.76%) |
|  | Hispanic | 2449<br>(19.71%) | 1718 (19.91%) | 602 (18.73%) | 129 (22.20%) |
| Insurance Status, n (%) | Private | 3209<br>(25.83%) | 2291 (26.55%) | 811 (25.23%) | 107 (18.42%) |
|  | Medicare | 6033<br>(48.56%) | 4033 (46.73%) | 1647 (51.24%) | 353 (60.76%) |
|  | Medicaid | 794<br>(6.39%) | 553 (6.41%) | 210 (6.53%) | 31 (5.34%) |
|  | Self/No insurance | 2389<br>(19.23%) | 1753 (20.31%) | 546 (16.99%) | 90 (15.49%) |

| Variables | Overall<br>n=12425 | ICH score 0-2<br>n=8630 | ICH score 3-4<br>n=3214 | ICH score 5-6<br>n=581 |
| --- | --- | --- | --- | --- |
| Smoking, n (%) | 1391<br>(11.20%) | 1081 (12.53%) | 275 (8.56%) | 35 (6.02%) |
| Drug/Alcohol Use, n (%) | 1216<br>(9.79%) | 889 (10.30%) | 291 (9.05%) | 36 (6.20%) |
| Hypertension, n (%) | 9469<br>(76.21%) | 6609 (76.58%) | 2398 (74.61%) | 462 (79.52%) |
| Diabetes, n (%) | 3076<br>(24.76%) | 2145 (24.86%) | 802 (24.95%) | 129 (22.20%) |
| Obesity, n (%) | 3005<br>(24.19%) | 2179 (25.25%) | 713 (22.18%) | 113 (19.45%) |
| Atrial fibrillation/Atrial flutter, n (%) | 1961<br>(15.78%) | 1271 (14.73%) | 574 (17.86%) | 116 (19.97%) |
| Coronary Artery Disease, n (%) | 1851<br>(14.90%) | 1229 (14.24%) | 507 (15.77%) | 115 (19.79%) |
| Peripheral Vascular Disease, n (%) | 364<br>(2.93%) | 253 (2.93%) | 96 (2.99%) | 15 (2.58%) |
| Prior Stroke or TIA, n (%) | 2862<br>(23.03%) | 1965 (22.77%) | 771 (23.99%) | 126 (21.69%) |
| Stroke Center Type, n (%) |  |  |  |  |
| CSC | 10458<br>(84.17%) | 7251 (84.02%) | 2711 (84.35%) | 496 (85.37%) |
| PSC | 1384<br>(11.14%) | 988 (11.45%) | 344 (10.70%) | 52 (8.95%) |
| TSC | 583<br>(4.69%) | 391 (4.53%) | 159 (4.95%) | 33 (5.68%) |
| Onset of Symptoms to Arrival Time in minutes, median (IQR) | 177 (61, 515) | 182.00 (62.00, 541.00) | 162.00 (60.00, 454.00) | 149.00 (60.00, 453.00) |
| Door to CT Time in minutes, median (IQR) | 19 (10, 52) | 18 (10, 62) | 21 (11, 43) | 26 (15, 46) |
| Region, n (%) | South | 5372<br>(43.24%) | 3732 (43.24%) | 1351 (42.03%) | 289 (49.74%) |
|  | West Central | 2454<br>(19.75%) | 1652 (19.14%) | 680 (21.16%) | 122 (21.00%) |
|  | East Central | 1568<br>(12.62%) | 1125 (13.04%) | 385 (11.98%) | 58 (9.98%) |
|  | North | 2340<br>(18.83%) | 1613 (18.69%) | 628 (19.54%) | 99 (17.04%) |
|  | Panhandle | 691<br>(5.56%) | 508 (5.89%) | 170 (5.29%) | 13 (2.24%) |
| Impaired Level of<br>Consciousness, n (%) |  | 3915<br>(31.51%) | 1917 (22.21%) | 1661 (51.68%) | 337 (58.00%) |
| In-hospital Mortality,<br>n (%) |  | 2129<br>(17.13%) | 487 (5.64%) | 1257 (39.11%) | 385 (66.27%) |
| Hospital Length of<br>Stay in days, median<br>(IQR) |  | 6.24 (3.17,<br>12.06) | 6.84 (3.92,<br>12.04) | 5.45 (2.03,<br>13.84) | 1.89 (0.95, 4.13) |
| Withdrawal of Life<br>Sustaining<br>Treatment, n (%) |  | 3393<br>(27.31%) | 1097 (12.71%) | 1873 (58.28%) | 423 (72.81%) |
| Discharge<br>Disposition, n (%) | Home/Rehab | 5759<br>(46.35%) | 5324 (61.69%) | 414 (12.88%) | 21 (3.61%) |
|  | Other | 6666<br>(53.65%) | 3306 (38.31%) | 2800 (87.12%) | 560 (96.39%) |

ICH indicates Intracerebral hemorrhage; WLST, withdrawal of life-sustaining treatment; TIA, Transient Ischemic Attack; CSC, Comprehensive Stroke Center; PSC, Primary Stroke Center; TSC, Thrombectomy-Capable Stroke Center; GCS, Glasgow Come Scale.
Data are presented as n (%) or median (Q1-Q3).

A total of 8631 (69%) of patients had an ICH score of 0-2, 3214 (26%) patients had ICH score 3-4, and 581 (5%) of patients had ICH score of 5-6. Overall, in-hospital mortality was observed in 2129 (17%) of all patients, with an average hospital length of stay of 6.25 days (SD = 14). In-hospital mortality rate for ICH group 0-2 was 6.6%, 41.5% for group 3-4 and 66% for group 5-6.

WLST decision was seen in 3393 (27%) patients overall.

### Model Performance for Predicting WLST Decision

A total of 12,425 individuals with documented ICH scores and outcome of WLST were included in the analysis. The Mean Decrease Gini metric was used to rank variable importance. The most predictive factors associated with WLST decision were ICH score value, age, region, impaired level of consciousness on presentation, insurance status, race, surgical treatment type, sex, stroke center type and diabetes. The variable importance plot highlighting the predictors most strongly associated with WLST decision can be seen in Figure 1. The most predictive factor associated with WLST decision was ICH score value. The RF and LR models exhibited strong discrimination in predicting WLST decision (RF model: AUC = 0.94; LR model: AUC = 0.82). ROCs are shown in Figure 2.

**Figure 1.**
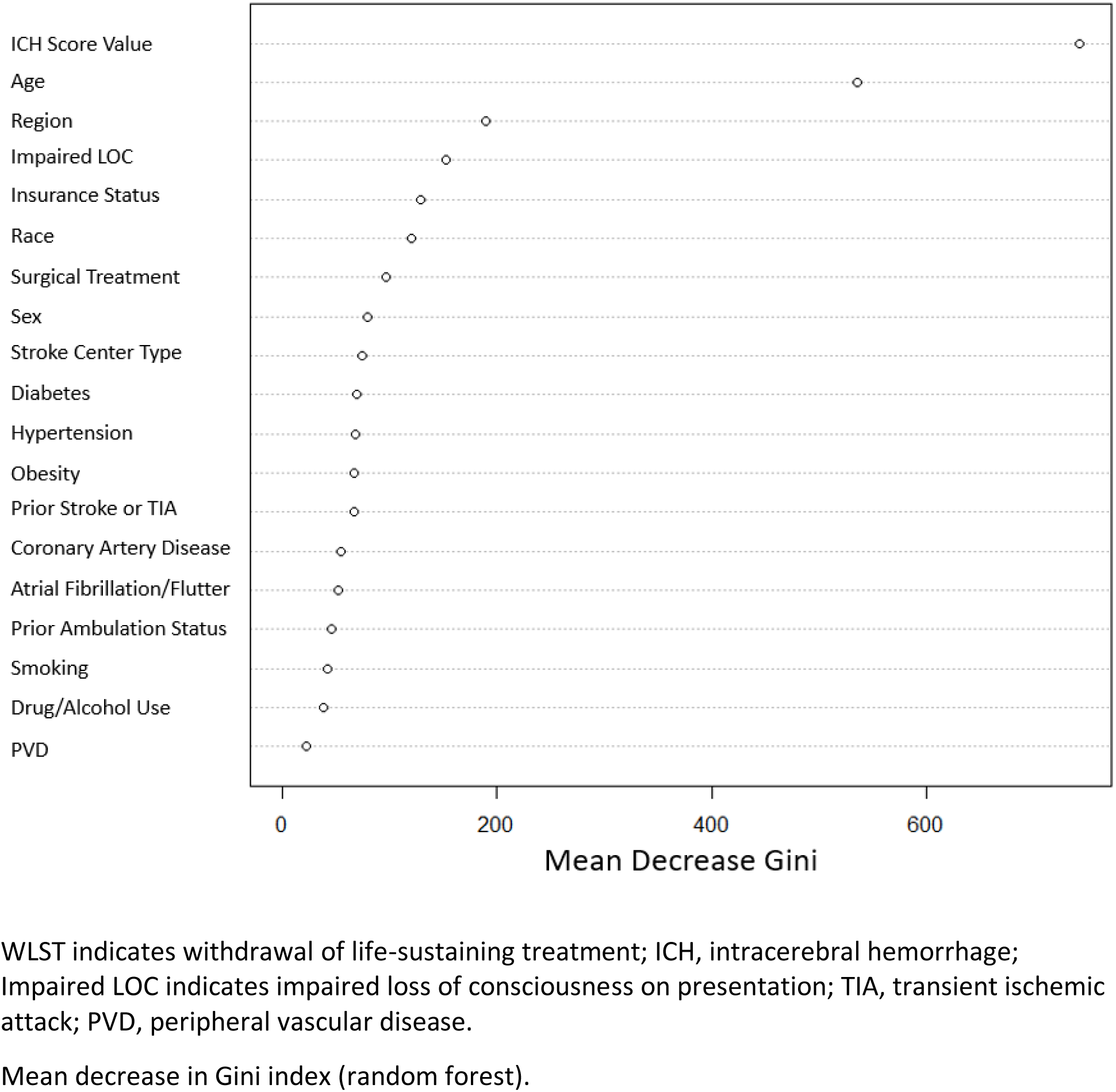
Variable Importance Plot for Predictors of WLST Decision in Random Forest Model.

**Figure 2.**
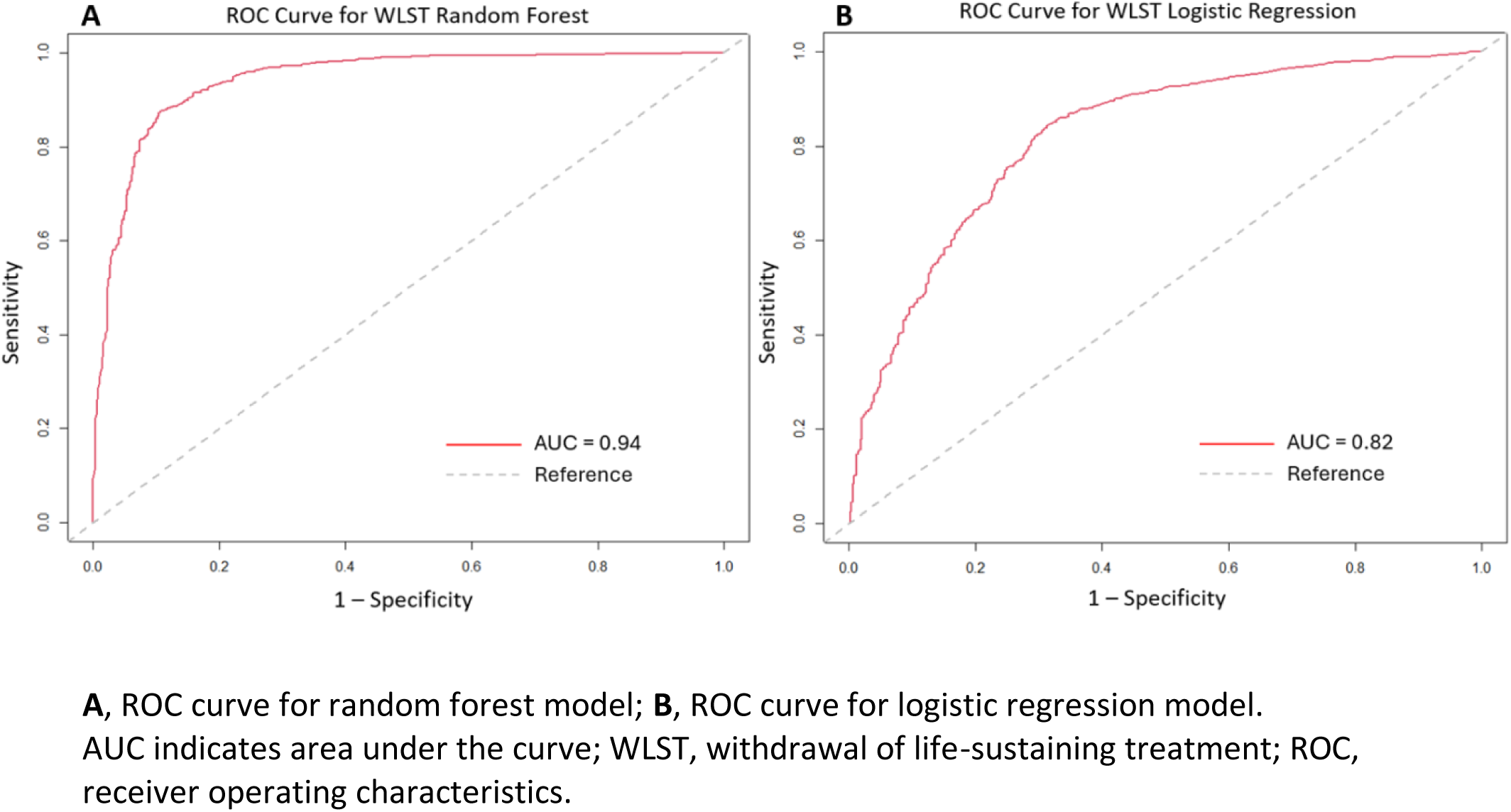
Receiver Operator Characteristic (ROC) Curves for Predictors of WLST Decision.

### Mortality and WLST Decision by ICH Score Group

Analysis of secondary outcomes was performed on data from 2015 to 2022 given a relatively low number of patient cases for the years 2013 and 2014. Univariate logistic regression analyses revealed a significant association between higher ICH scores and increased odds of mortality (Table 2). Compared to patients with an ICH score of 0-2, those with a score of 3-4 had an odds ratio (OR) of mortality of 10.69 (95% CI: 9.49-12.04, p < .0001), while those with a score of 5-6 had an OR of 34.40 (95% CI: 28.14-42.04, p < .0001). With regards to WLST, patients with ICH scores of 3-4 had an OR of 9.35 (95% CI: 8.50-10.30, p < .0001) for WLST decision, while for scores of 5-6, the OR was 18.64 (95% CI: 15.28-22.74, p < .0001), compared to the reference group. The likelihood of early WLST (occurring within the first two days after presentation) was also associated with higher ICH scores. Patients with an ICH score of 3-4 had an OR of 2.97 (95% CI: 2.48-3.55, p < .0001) for early WLST, while those with an ICH score of 5-6 had an OR of early WLST of 9.51 (95% CI: 7.33-12.35, p < .0001).

**Table 2.** Univariate Logistic Regression for Mortality and WLST by ICH Score Group and Time Period, 2015 to 2022.

| Outcome | ICH Score Group | Overall |  | 2015-2018 |  | 2019-2022 |  |
| --- | --- | --- | --- | --- | --- | --- | --- |
|  |  | OR | 95% CI | OR | 95% CI | OR | 95% CI |
| Mortality | 0-2 | Reference | - | Reference | - | Reference | - |
|  | 3-4 | 10.69 | 9.49-12.04 | 11.90 | 9.67-14.65 | 10.11 | 8.75-11.68 |
|  | 5-6 | 34.40 | 28.14-42.04 | 46.85 | 32.05-68.47 | 30.24 | 23.82-38.38 |
| WLST | 0-2 | Reference | - | Reference | - | Reference | - |
|  | 3-4 | 9.35 | 8.50-10.30 | 9.90 | 8.28-11.84 | 9.30 | 8.29-10.43 |
|  | 5-6 | 18.64 | 15.28-22.74 | 16.25 | 11.61-22.76 | 20.67 | 16.10-26.53 |
| Early WLST | 0-2 | Reference | - | Reference | - | Reference | - |
|  | 3-4 | 2.97 | 2.48-3.55 | 2.98 | 2.11-4.22 | 2.95 | 2.39-3.63 |
|  | 5-6 | 9.51 | 7.33-12.35 | 10.08 | 6.07-16.75 | 9.28 | 6.84-12.58 |

ICH indicates Intracerebral Hemorrhage; WLST, Withdrawal of Life-Sustaining Treatment. Early WLST refers to WLST decision made within day 0 or 1 after hospital presentation.
The reference group for all outcomes is ICH score 0-2, which is assigned as an odds ratio of 1.0. Models were adjusted for age, sex, race/ethnicity, comorbidities (hypertension, diabetes, obesity, prior stroke or TIA), region, insurance status and hospital type.
All ORs for each ICH score group and time period were statistically significant with $p < 0.0001$

### Temporal Differences in Mortality by ICH Score Group

Stratified univariate logistic regression was performed for times periods 2015-2018 and 2019-2022. In the 2015–2018 cohort, patients with ICH scores of 3–4 had an OR of 11.90 (95% CI: 9.67–14.65, p < 0.0001) for mortality, while those with ICH scores of 5–6 had an OR of 46.85 (95% CI: 32.05–68.47, p < 0.0001) compared to the reference group (ICH score 0–2). In the 2019–2022 cohort, patients with ICH score 3-4 had an OR of 10.11 (95% CI: 8.75–11.68, p < 0.0001) for mortality, while those with ICH scores of 5-6 had an OR of 30.24 (95% CI: 23.82– 38.38, p < 0.0001) for mortality (Table 2). Temporal trend in mortality by ICH Score group can be seen in Figure 3A.

**Figure 3:**
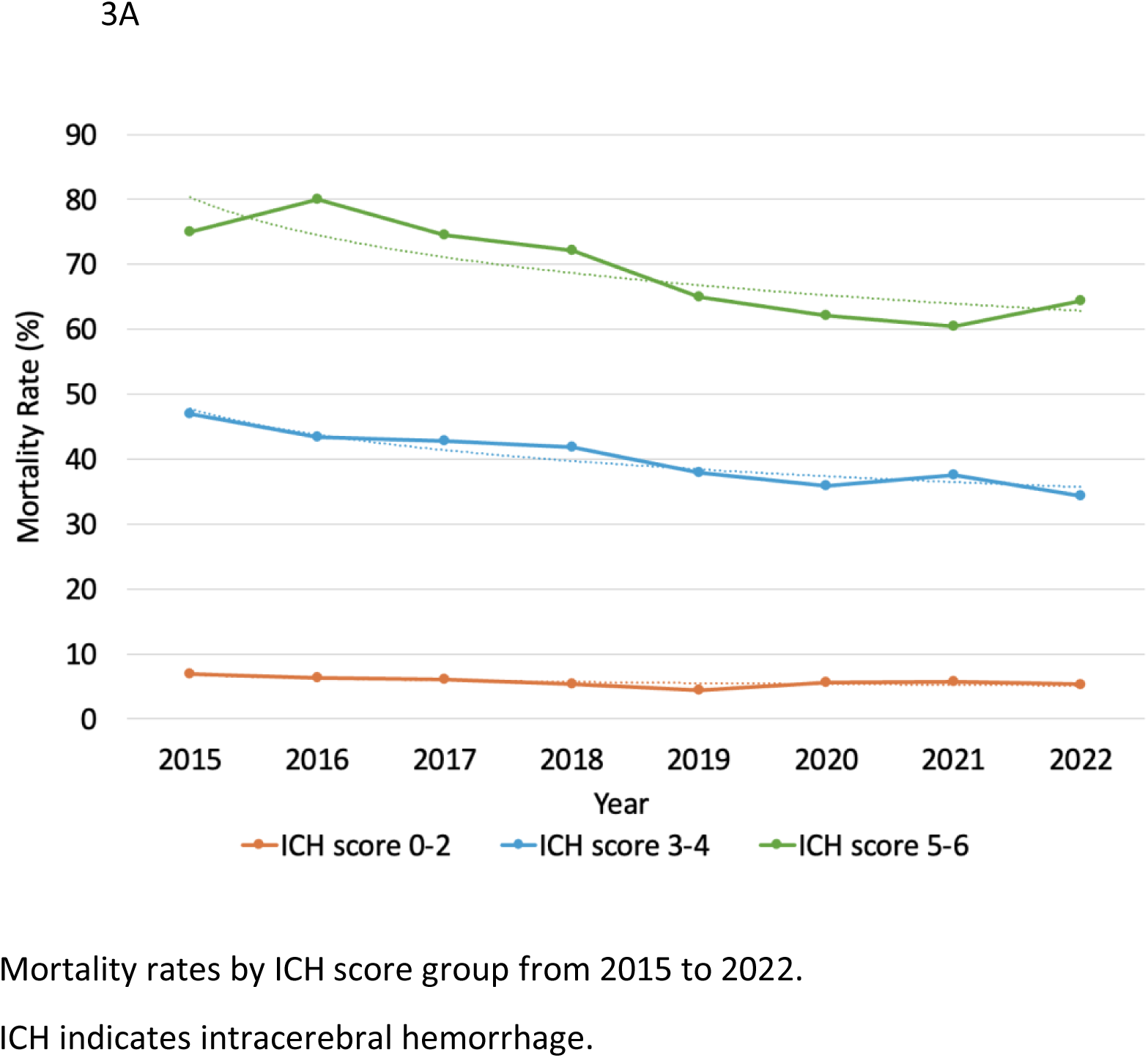
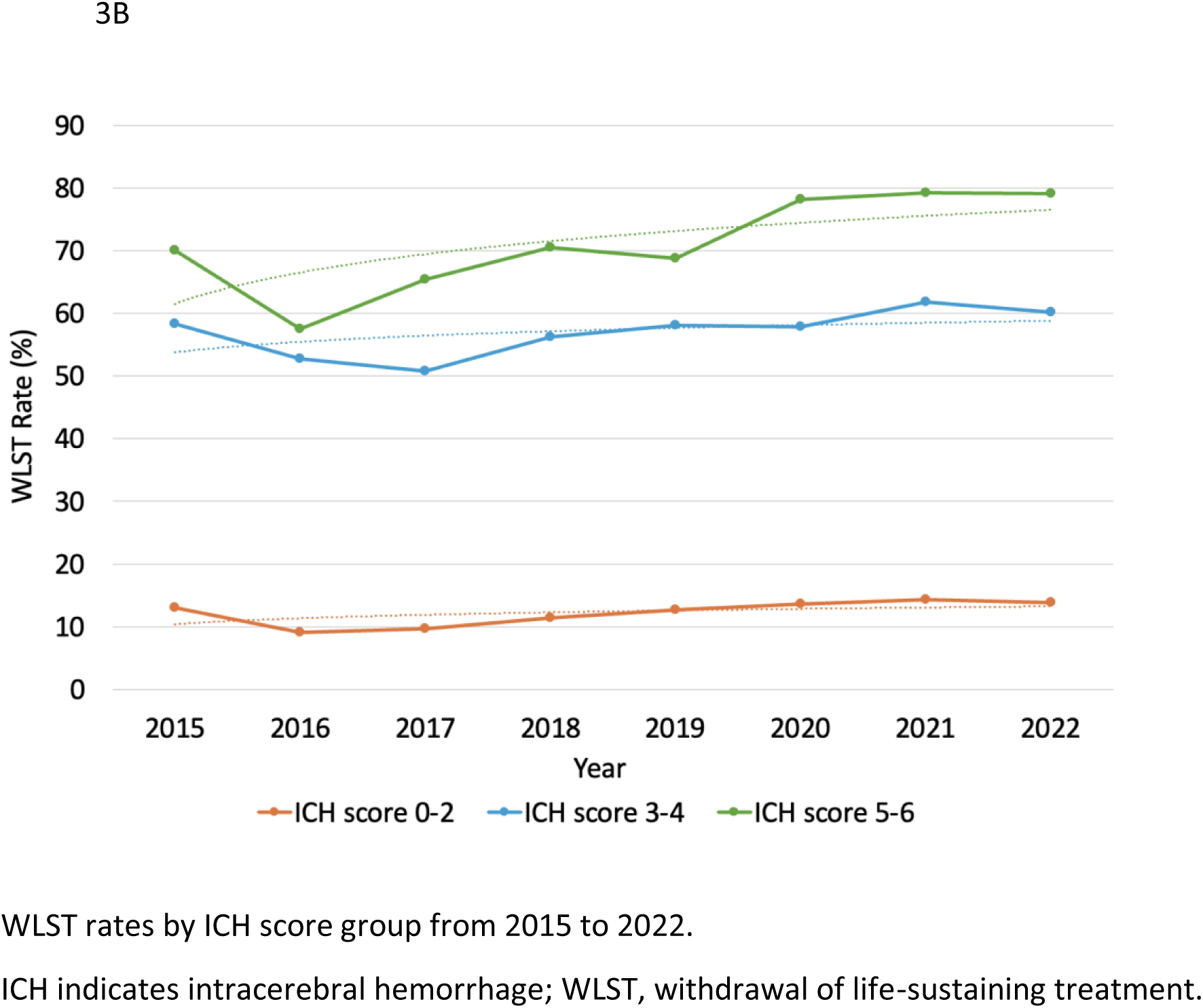
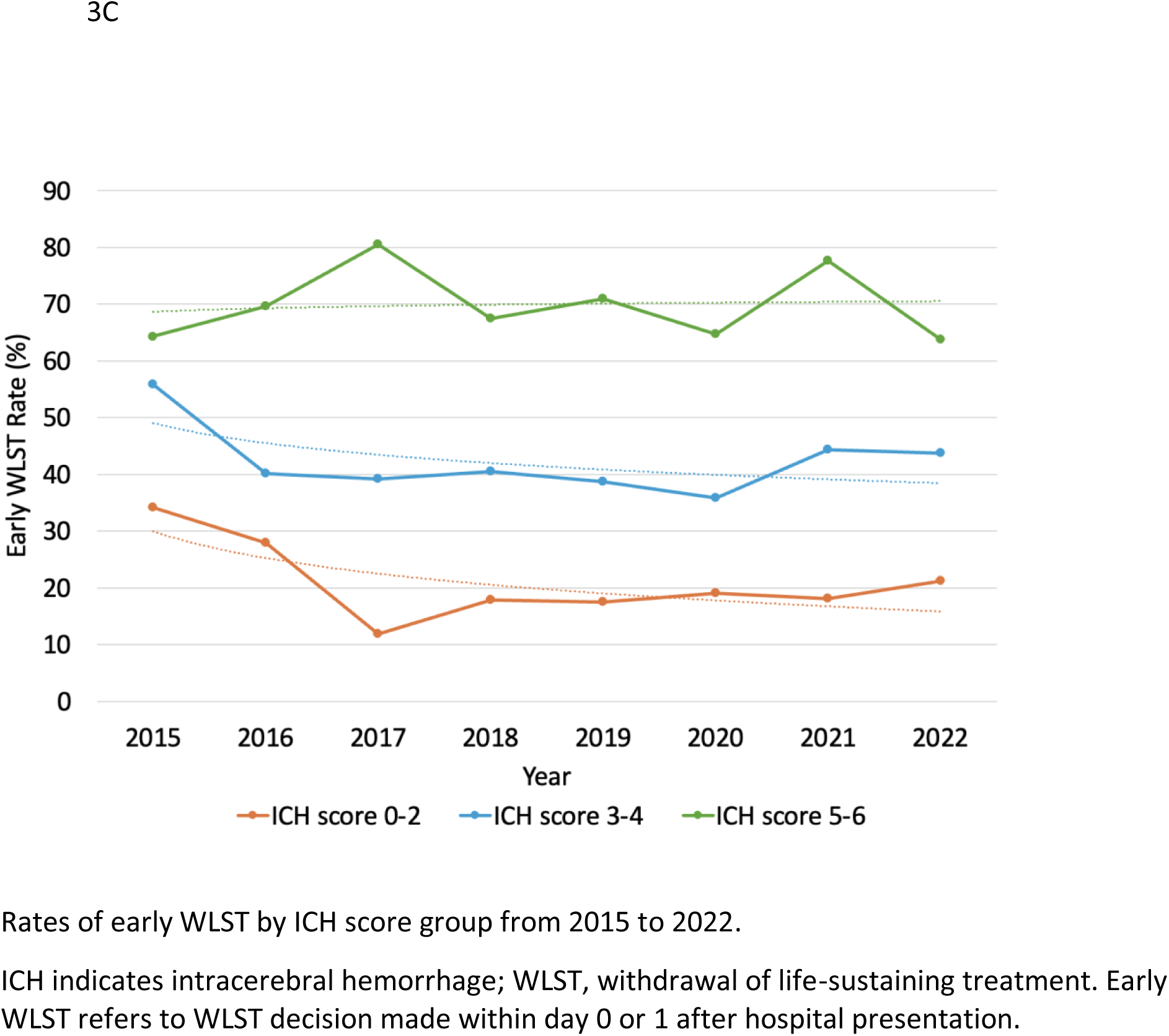
Temporal Trends in Mortality and WLST by ICH Score Group

### Temporal Differences in WLST by ICH Score Group

Higher ICH scores were strongly associated with increased odds of WLST across both time periods. In the 2015–2018 cohort, OR of WLST was 9.9 (95% CI: 8.28–11.84, p < 0.0001) for the ICH score 3-4 group and 16.25 (95% CI: 11.61–22.76, p < 0.0001) for the ICH score 5-6 group. Comparatively, in the 2019–2022 cohort, odds of WLST were 9.3 (95% CI: 8.29–10.43, p < 0.0001) and 20.67 (95% CI: 16.10–26.53, p < 0.0001) for ICH score 3–4 and 5–6 groups, respectively.

There was also a significant association in both time periods between ICH scores and early WLST specifically (0 to 1 day after presentation). Patients with ICH scores of 3–4 in the 2015-2018 cohort had an OR of 2.98 (95% CI: 2.11–4.22, p < 0.0001), while those with ICH scores of 5–6 had an OR of 10.08 (95% CI: 6.07–16.75, p < 0.0001). For the 2019–2022 cohort, the odds of early WLST were 2.95 (95% CI: 2.39–3.63, p < 0.0001) for the ICH score 3–4 group and 9.28 (95% CI: 6.84–12.58, p < 0.0001) for the ICH score 5–6 group (Table 2). Temporal trends in both WLST and early WLST by ICH Score group can be seen in Figures 3B-C.

## DISCUSSION

In this prospective, large, multicenter stroke registry of patients with ICH between 2013 and 2022, there was an overall mortality rate of 17%. This mortality rate was largely driven by the decision to withdrawal life-sustaining treatment; WLST occurred in 27% of all patients. Investigating mortality by score value, patients with admission ICH score of 5 to 6 had a mortality rate of 69%, ICH score 0 to 2 had a mortality rate of about 6%, and group with ICH scores from 3 to 4 had 40% in-hospital mortality overall. Higher ICH score value appeared to be strongly associated with mortality. This association between ICH score and increased odds of mortality appeared to be similar comparing 2015-2018 to 2019-2022 time periods; for the higher ICH score value (5 or 6) group, the odds of mortality was lower in 2019 compared to 2015-2018.

In this study, we identified the most predictive variable associated with WLST decision to be ICH score severity. Of note, other factors we identified as being associated with WLST included age, region within the state, level of consciousness on presentation, insurance status and race.

Recently, socioeconomic status and other social determinants of health have been noted to be associated with WLST after ICH.^11^ Region, insurance status, and race being notable variables influencing WLST in our cohort appears to again demonstrate this association. Nevertheless, the factor with the highest importance in predicting WLST decision across our cohort was the ICH score.

Compared to patients with lower documented ICH scores (0 to 2), patients in the higher ICH score groups were associated with higher likelihood of decision to WLST. For patients with ICH score value of 3 or 4, this likelihood was consistent across the time periods. There did appear to be a stronger association between decision to WLST and ICH score 5 to 6 in 2019-2022 than there was in 2015-2018. Furthermore, the chances of early decision to WLST did appear to be lower than overall WLST, however there is still an association between ICH score group and likelihood of early WLST. This appeared consistent across the time periods.

The ICH score grading scale was proposed to provide information and assess treatment benefits and risks for patient families, rather to utilizing the score for prognosticating outcomes.^10,12^ Since the initial publication of the grading scale, there has been a tendency by healthcare providers to use this scale as a prognostication tool to influence clinical decision-making.^8,13,14^ Like other clinical grading scales for disease entities within neurocritical care, ICH score value appears to influence outcomes, leading to a self-fulfilling prophecy whereby aggressive treatment of patients in traditionally high severity groups is deemed futile. This highly influences perception by the medical team along with communication with patient families, leading to decision to withdraw life sustaining treatment.^4,15–17^

Intracerebral hemorrhage is associated with high morbidity and mortality, and in addition often with higher rates of WLST than other disease processes within neurocritical care, making it difficult to disentangle mortality from the influence of care decision-making.^16–19^ Recent data from our large multicenter FSR noted rates of WLST to be 9% in acute ischemic stroke, 28% in ICH and 19% in subarachnoid hemorrhage between 2008 and 2021.^2^ Given the nihilism surrounding prognosis in patients with ICH historically, it is not surprising that we identified the most predictive variable associated with WLST to be initial ICH score value.

As this study explored association with ICH score value and temporal differences in practice with respect to WLST, it is important to consider relevant guideline recommendations surrounding the study period. American Heart Association/American Stroke Association (AHA/ASA) management guidelines published in 2010 highlight the uncertainty and potential for self-fulfilling prophecies of poor outcome with use of ICH score.^20^ At the time, it was suggested that physicians avoid recommendations of care limitations such as do not resuscitate (DNR) orders or WLST within at least the second full day of hospitalization.^20–22^

A study from 2015 observed that patients without placement of do not resuscitate (DNR) orders within the first 5 days after ICH presentation had a substantially lower 30-day mortality than predicted by the ICH score.^23^ That same year, AHA/ASA practice guidelines were updated and added to their recommendations that in patients without preexisting advanced directives specify as such, that postponing DNR or WLST until at least the second full day of hospitalization is reasonable to improve function outcome and mortality in spontaneous ICH.^24^ In addition, that DNR orders should also not limit appropriate medical and surgical interventions in ICH in the first few days after ICH.^25^ Most recent guidelines continue to make these practice recommendations.^26,27^

Revisiting our study findings from the FSR and relevant practice guidelines, despite these recommendations made during the corresponding time frames, we noted a continued association between higher ICH scores and decision to WLST. While only a proportion of WLST does appear to occur within the first two days of hospital admission, we found a persistent association between ICH score value and early WLST, without a notable change between the two time periods in the study.

We also observed a continued association between higher ICH score and likelihood of mortality across the study time periods. Mortality did appear to be lower in the 2019-2022 time period for the highest severity group (ICH score 5-6), despite WLST occurring more for that ICH score group within that time period. Improvement in mortality over time is not surprising given efforts over time to improve and bundle care with respect to prompt blood pressure lowering, anticoagulant reversal, improved access to neurosurgery, among other efforts.^28^ Even more recently, new evidence suggests surgical techniques such as minimally invasive hematoma evacuation is associated with improvement in mortality and functional outcome. ^29,30^

There may have been an influence of the COVID-19 pandemic in-hospital mortality over the 2019 to 2022 time period, potentially resulting in higher mortality rates than there would have been in the absence of a global health crisis.^31–33^ Moreover, resource limitations and potential comorbidities during that time could have influenced WLST decision-making during the pandemic, resulting in this increase the highest severity group.^34^ Data regarding COVID-19 testing and rates of comorbid infection in our patients could have helped elucidate this, however this information was not available. Nonetheless, assuming a continued trajectory toward more effective therapeutics for ICH, we anticipate not only a continued trend toward decreased in-hospital mortality, but in addition a potential decrease in WLST in favor of more aggressive approach to patient treatment over time.

Decisions regarding WLST have both hindered and influenced understanding of recovery in this disease process.^19^ There is a paucity of high-quality data investigating outcomes in ICH beyond 3 months, and even less beyond 6 months following initial presentation. ^9,35,36^ In a large cohort of over 500 ICH patients from a prospective registry over 5 years of patients who were maximally treated, there was a long-term mortality rate of 30%, and 45% of patients reached favorable functional outcome by modified Rankin Scale (mRS) 0 to 3 at 12-month follow up, suggesting a significant proportion of the patients had meaningful neurologic improvement.^8^ A subsequent multicenter validation study compared outcome prognostication of the max-ICH score, which excludes patients without early care limitations, to that of the ICH score, and demonstrated improved prognostication of functional outcome for ICH patients without early care limitations.^13^ Less still has been published regarding 12-month outcome trajectories in ICH patients with severe disability specifically; one such study demonstrated that among those with mRS of 4 to 5 at 30-day following ICH, over 40% recovered to favorable functional outcome (mRS 0-3) at 1 year.^37^

There are limitations with respect to our study. Firstly, important relevant information regarding code status, prior advanced directives, extent to which medications and medical management was continued or withheld is not available in the FSR and therefore unable to be considered in this study. In addition, details regarding WLST decision and extent to which this was influenced by patient and family-centered views was also not available. Within the FSR, ICH score had high missingness, although 12,426 had documented scores and patient characteristics were similar to those without ICH score. Additionally, the FSR does not include data on cognitive and functional outcomes after hospitalization, making it difficult to fully assess the impact of the self-fulfilling prophecy in this population over time.

Withdrawal of life sustaining treatment in ICH continues to be a frequent occurrence due to concerns about futility with continued aggressive management, particularly for patients with high ICH scores. Despite efforts in the field to counsel providers away from early WLST particularly based on ICH score severity, ICH score continues to be the most predictive variable associated with WLST, and decision to withdraw life-sustaining therapy with the first two days of presentation also continues to occur. Our findings suggested a continued influence of the self-fulfilling prophecy in ICH.

## ACKNOWLEDGEMENTS

Drs. Massad, Alkhachroum, Asdaghi and Gardener were involved in the study concept and design. Drs. Zhou, Ying, Gutierrez and Gardener were involved in the acquisition of data. Drs. Zhou, Manolovitz and Gardener were involved in the statistical analysis. Drs. Zhou, Manolovitz, Massad, Alkhachroum and Gardener were involved in analysis and interpretation of data. Drs. Massad, Alkhachroum, Zhou, Manolovitz, Asdaghi, Gardener, Ying, Gutierrez, Jameson, Rose, Kottapally, Merenda, O’Phelan, Koch, Romano and Rundek were involved in drafting the article. Drs. Massad, Alkhachroum, Gutierrez and Asdaghi were involved in study supervision.

